# COVID-19 in England: spatial patterns and regional outbreaks

**DOI:** 10.1101/2020.05.15.20102715

**Authors:** Claudio Fronterre, Jonathan M Read, Barry Rowlingson, Simon Alderton, Jessica Bridgen, Peter J Diggle, Chris P Jewell

## Abstract

**Aims:** to investigate the spatiotemporal distribution of COVID-19 cases in England; to provide spatial quantification of risk at a high resolution; to provide information for prospective antigen and serological testing.

**Approach:** We fit a spatiotemporal Negative Binomial generalised linear model to Public Health England SARS-CoV-2 testing data at the Lower Tier Local Authority region level. We assume an order-1 autoregressive model for case progression within regions, coupling discrete spatial units via observed commuting data and time-varying measures of traffic flow. We fit the model via maximum likelihood estimation in order to calculate region-specific risk of ongoing transmission, as well as measuring regional uncertainty in incidence.

**Results:** We detect marked heterogeneity across England in COVID-19 incidence, not only in raw estimated incidence, but in the characteristics of within-region and between-region dynamics of PHE testing data. There is evidence for a spatially diverse set of regions having a higher daily increase of cases than others, having accounted for current case numbers, population size, and human mobility. Uncertainty in model estimates is generally greater in rural regions.

**Conclusions:** A wide range of spatial heterogeneity in COVID-19 epidemic distribution and infection rate exists in England currently. Future work should incorporate fine-scaled demographic and health covariates, with continued improvement in spatially-detailed case reporting data. The method described here may be used to measure heterogeneity in real-time as behavioural and social interventions are relaxed, serving to identify “hotspots” of resurgent cases occurring in diverse areas of the country, and triggering locally-intensive surveillance and interventions as needed.

**Caveats:** There is general concern over the ability of PHE testing data to capture the true prevalence of infection within the population, though this approach is designed to provide measures of spatial prevalence based on testing that can be used to guide further future testing effort. Now-casts of epidemic characteristics are presented based on testing data alone (as opposed to “true” prevalence in any one area). The model used in this analysis is phenomenological for ease and speed of principled parameter inference; we choose the model which best fits the current spatial case timeseries, without attempting to enforce “SIR”-type epidemic dynamics.

## 1. Introduction

As the UK enters the next stage of the COVID-19 pandemic, and considers relaxation of current BSI (Behavioural and Social Interventions, i.e. “lockdown”), accurate estimation of disease prevalence at a fine spatial scale is vital for informing and underpinning proactive disease management. A common characteristic of previous infectious disease outbreaks, most notably in the West African Ebola outbreak 2013–2016, is that post-peak epidemics are typically characterised by small focal outbreaks of disease occurring in disparate regions at different times. Spatial heterogeneity in incidence was observed in the UK for pandemic influenza, and the COVID-19 epidemic in the UK is exhibiting spatial heterogeneities. The ability to detect such spatial outbreaks is therefore key to efficient local containment, preventing local foci of infection degenerating into a nationally resurgent epidemic wave.

In determining a suitably “fine” spatial scale at which to perform surveillance, report the occurrence of disease cases, and be able to predict risk from continued outbreaks, it is necessary to consider the spatial variation of underlying population characteristics determining transmission of, and susceptibility to, COVID-19. For example, clinical studies have highlighted the importance of comorbidities, such as obesity and diabetes, and socioeconomic and ethnic background on severity of known to vary with space, even over short distances. Such potential risk factors are routinely collected COVID-19 infection [Docherty *et al*., 2020; Fang *et al* 2020; Pareek *et al*., 2020], factors which are known to vary with space, even over short distances. Such potential risk factors are routinely collected via Census information at geographically small scales, such as Lower Super Output Area. Thus there is a pressing need for reliable COVID-19 case reporting data at a spatial resolution to match underlying covariate data.

In this paper, we analyse COVID-19 data since April 1st 2020 in England, disaggregating PHE-reported positive test cases at the Lower Tier Local Authority (LTLA). Our aim is to detect regions that have unusually high incidence of disease relative to the underlying population, and distinguish regions with high potential for self-sustaining transmission from those at risk of importing infection from other regions. We use established spatio-temporal methodology [Held *et al*., 2005] to outline an approach for providing now-casts of the spatial outbreak characteristics on a daily time scale, provided that reliable data are available at a high spatial resolution. Finally, we discuss these results in the context of spatial analysis of data from the “COVID Symptom Study” (https://covid.joinzoe.com/) which provides a far greater level of spatial granularity in self-reported COVID-19 symptom prevalence.

## 2. Methods

In this section, we describe a spatiotemporal phenomenological approach to monitoring COVID-19 on a national scale, highlighting regions of higher than expected case incidence, regions that have high propensity for sustained transmission, and regions that are at risk from imported infection from other regions of the country. We begin by describing our various data sources, before describing the analytic model and fitting process.

### Data

#### Case Data

We base our analysis on positive test results in England, as reported by Public Health England (PHE, https://coronavirus.data.gov.uk/). Although these data may be subject to temporal biases due to changing testing regimes, they appear to provide the most spatially resolved measure of number of COVID-19 cases available to modellers with cases attributed to each of 315 Lower Tier Local Authorities (LTLAs) in England consistent with our aim of spatial analysis of the outbreak (modified from statutory LTLAs, see following section). As of 8th May 2020, these data contain 129320 cases attributed to a LTLA. The overall case time series is shown in Figure 1, exhibiting a decline from early April, a strong weekend effect, and also evidence of a 4 day lag in reporting. The spatial weekly case prevalence (number of cases divided by population size) for 6th April to 3rd May (discounting the latest 4 observations up to 7th May as unreliable) inclusive in Figure 2 shows marked variation over England.

**Figure 1:**
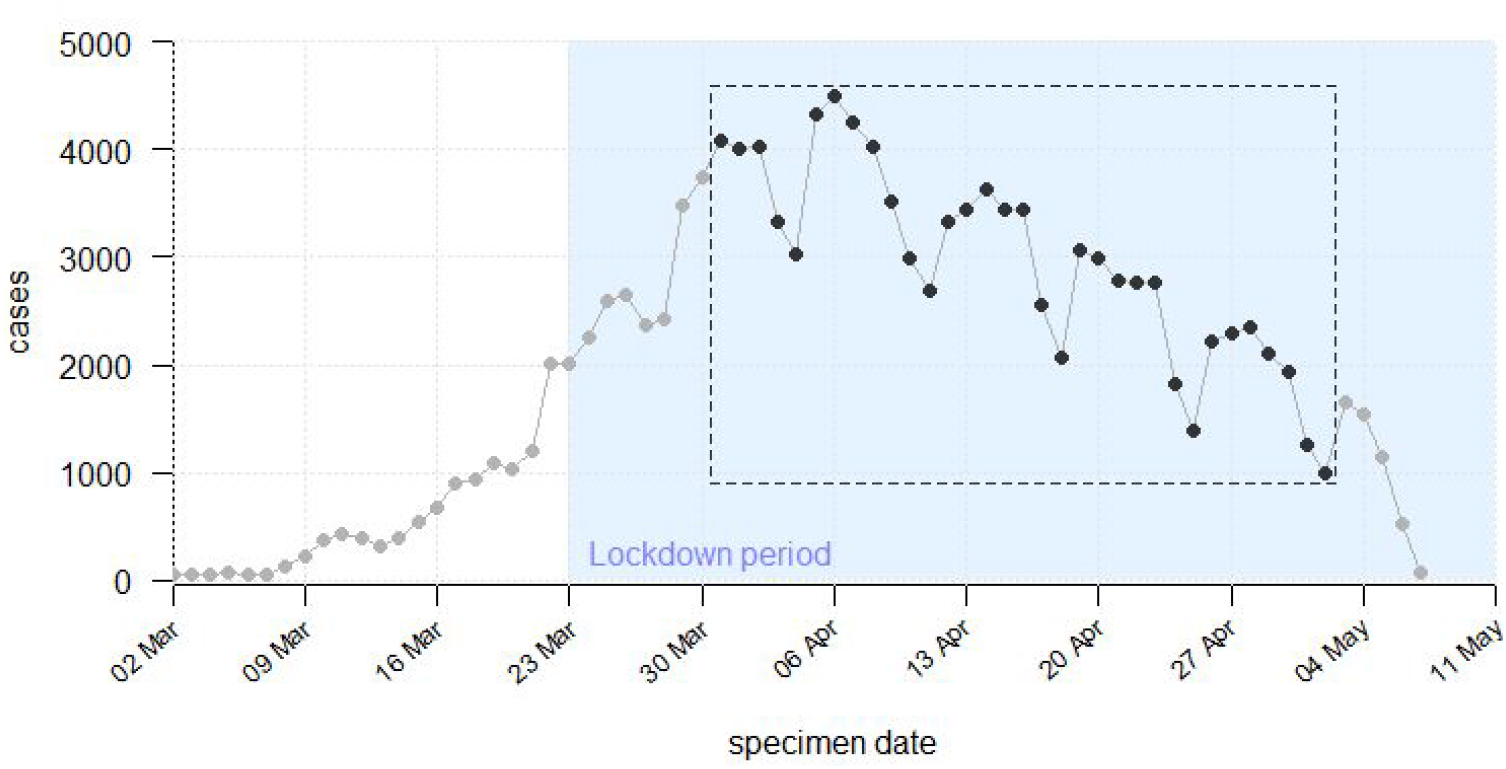
COVID-19 case time series up to 7th May 2020, England. Data from 1st April to 3rd May only is analysed, denoted by the dashed box.

**Figure 2:**
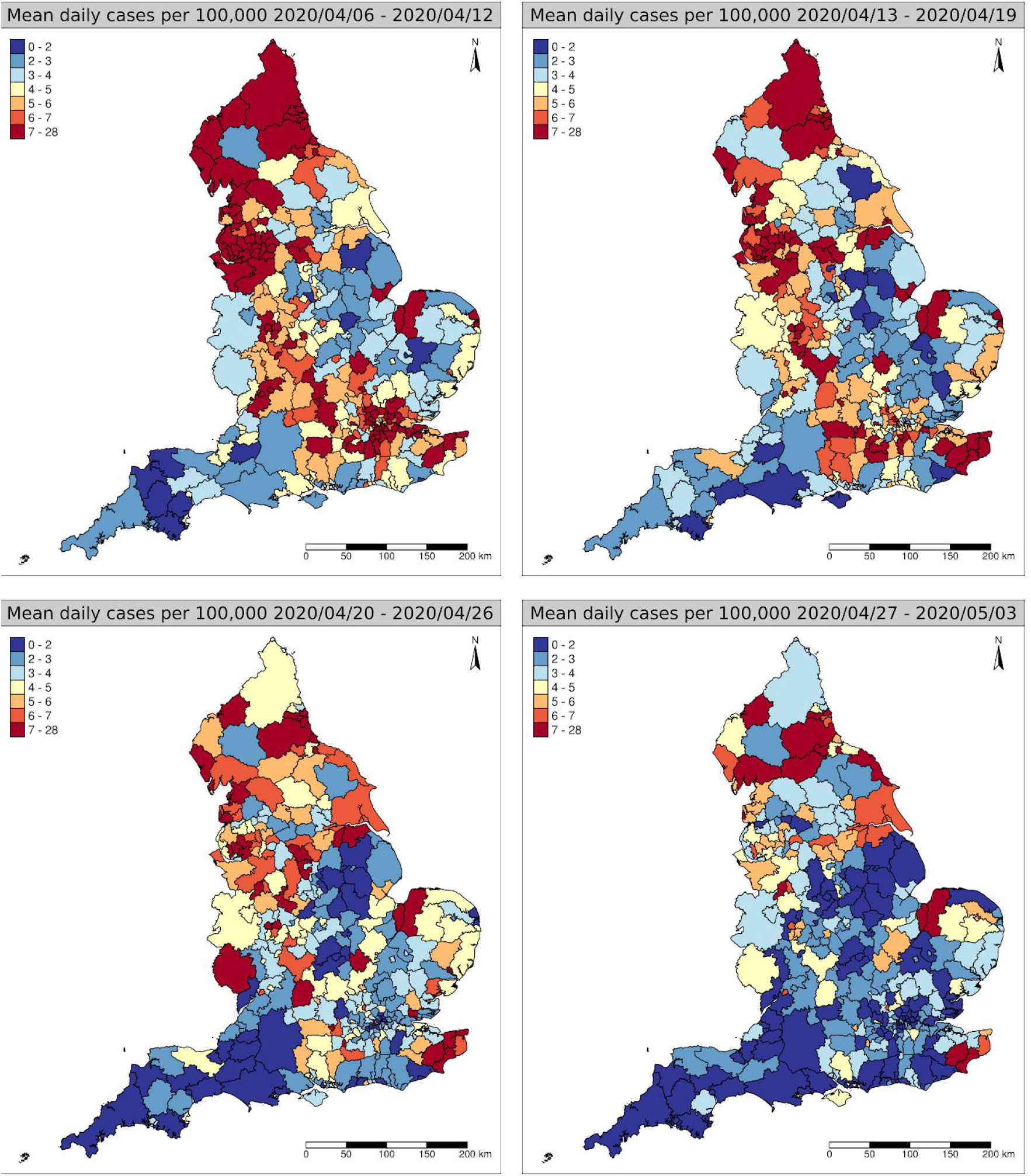
Spatial distribution of mean daily incidence per 100,000 people in England by LTLA for weeks 8th April –– 3rd May inclusive.

Given the obvious change in overall epidemic dynamics after 23rd March (when BSI were implemented nationally), we henceforth perform our analysis on the time series from 1st April to 3rd May, discounting the 4 latest days’ observations as biased due to reporting lag.

#### Inter-LTLA connectivity

To establish connectivity between LTLAs in England, Census 2011 commuting volume data was aggregated from Middle Super Output Area (MSOA). This necessitated the aggregation of two pairs of LTLAs (Cornwall and Scilly, and City of Westminster and City of London) to allow mapping of MSOAs onto LTLAs. This reduced the statutory number of LTLAs from 317 to 315.

Census 2011 commuting information provides an estimate of the number of journeys made from “Residence” to “Workplace” MSOAs, which when aggregated to our LTLA mapping provides a matrix *W* of dimension 315×315. Importantly commutin *W* is non-symmetric, reflecting commuting behaviour rather than the reciprocity of disease transmission. We calculated a symmetric matrix *W*^★^ of the daily number of *journeys* between each LTLA

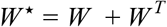

assuming that commuters return to their Residence each day, and go from their Residence to their Workplace and back at most once per day. The use of *W*^★^ was found to degrade model fit compared *W*, and was therefore discarded.

#### Traffic volume

Since inter-LTLA commuting data were derived from “business as usual” conditions in England, we assume that commuting is modulated during the COVID-19 outbreak by a relative measure of traffic flow provided by the UK Department for Transport (DfT). We construct a time series of traffic flow by taking DfT’s estimate of daily domestic car usage across the UK, expressed as a fraction of car usage on 1st February (Figure 3). The data used for our analysis covers the time period from 1st February 2020 to 1st May 2020 inclusive, and so were extended back to 1st January 2020 and forward to 7th May 2020 assuming values equal to their closest known neighbour in the overall time series.

**Figure 3:**
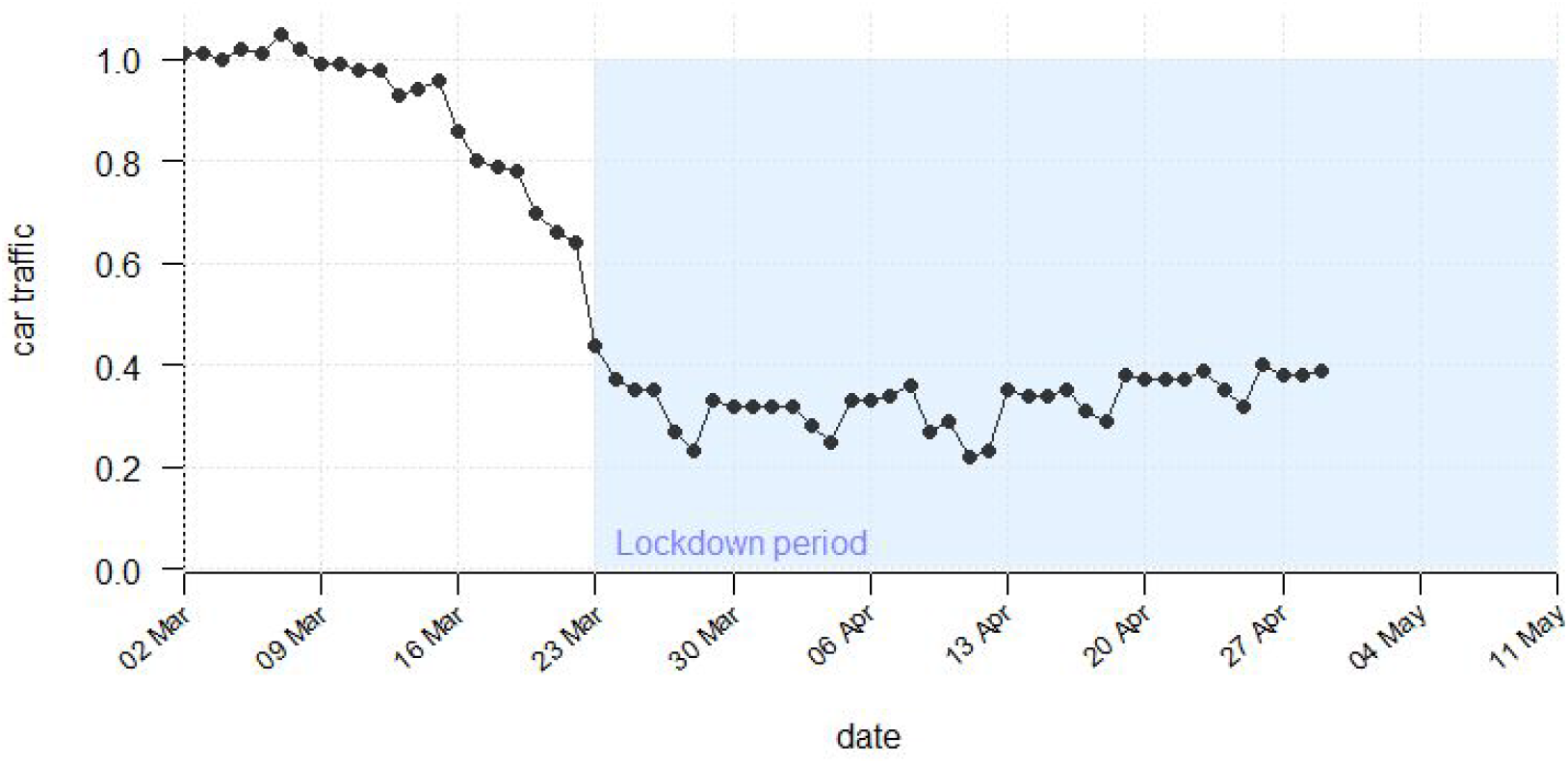
Road travel flow in England, expressed as a fraction of flow on 1st February 2020.

Potential covariates associated with the decline in cases are firstly the implementation of BSI on 23th March 2020, as well as observational information on traffic flow in England, retrieved from a national summary of domestic car travel (Figure 3).

#### Other Covariates

Other important covariates are the estimated total population size for each LTLA taken from ONS December 2019 predictions, together with a dummy variable encoding weekday vs weekend (to account for obvious weekend dips in case data).

#### Model

We model the progression of positive tests at the LTLA region level in England using a spatially coupled autoregressive time series. Let *Y_it_* be the number of positive COVID-19 tests in LTLA *_i_* on day *_t_* which we model as a negative binomial distributed random variable with mean and μ_*it*_ overdispersion *r*

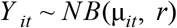

In each region, we model μ_*it*_ as the sum of a “intrinsic” time-homogeneous force of infection ε_*it*_, an autoregressive component within each region λ_*it*_ measuring the effect of within-region transmission, and a spatial autoregressive term ϕ_*it*_ measuring the effect of between-region transmission

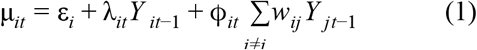

This makes the implicit assumption that the number of susceptible individuals does not change through time. Given the current low prevalence of disease – and therefore immunity – generally, we consider this to be a good approximation to the underlying epidemic process.

We decompose each component of transmission in turn. For the intrinsic transmission, we weight each region by the population size, such that

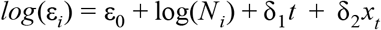

where *N_i_* is an offset representing the population size of LTLA _*i*_, and ε_0_ is the baseline intrinsic incidence. Furthermore, *t* represents time and *x_t_* an indicator for weekend days with associated coefficients δ.

For the within-region autoregressive term, we assume

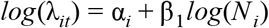

where α_*i*_ is a assumed to be a region-specific transmission rate, allowing some regions to be above or below the national average in terms of ongoing (i.e. autoregressive) infection risk, independently Normally distributed with mean λ*_0_* and variance τ*^2^*.

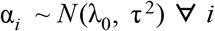

For the spatial force of infection term we assume

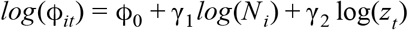

where ϕ_0_ is the mean spatial force of infection, *z_t_* is the observed England-wide traffic flow on day *_t_* expressed as a fraction of the flow on 1st February 2020. Importantly, the term *w_ij_* in Equation 1 is the mean daily number of commuters between regions *i* and *j*, allowing us to compute the sum of the force of infection on region *i* from all other regions *j* ≠ *i*.

#### Inference

Since we are interested in investigating current dynamics of the COVID-19 outbreak between LTLAs in England, we fit to “lockdown” case data only, taken to be from 1st April 2020 to allow for a 7 day delay since lockdown measures were imposed (23rd March), and up to 3rd May beyond which data are regarded as incomplete. Parameter inference is performed using maximum likelihood estimation in the R package “surveillance” (version 1.18, Meyer *et al*., 2017) and R version 3.6.3, allowing us to calculate the contribution to incidence in each region due to the intrinsic, within-region autoregressive, and between-region autoregressive terms. This allows us to distinguish between regions with unusually high incidence due to within-region epidemics, and those with unusually high transmission emanating from other areas. An advantage of this method is in its rapidity, with models fitting within minutes in order to provide real-time estimates of epidemic risk.

## 3. Results

Our results show a marked heterogeneity across England in COVID-19 incidence up to 3rd May 2020, not only in raw estimated incidence (Figure 2), but in the characteristics of within-region and between-region dynamics of PHE testing data. The choropleth maps in Figure 4 show the contribution to disease incidence of the within- and between-region autoregressive parts of the model and intrinsic component. The three components should be interpreted together. The Intrinsic component of the model establishes a baseline incidence across the whole of England, exhibiting a small linear decrease with respect to time (relative risk 0.978 (0.976, 0.980) per day, see Table S1). The Within- and Between-region maps then outline departures from this overall trend. The Within-region map identifies regions that are primarily driven by a local epidemic process, such that the outbreak may be considered still increasing in these regions. County Durham, King’s Lynn and West Norfolk, and Ashford are particularly affected, together with other regions of the North West. The Between-region map indicates regions in which incidence is primarily driven by importation of Within-region map identifies regions that are primarily driven by a local epidemic process, such that infection from other parts of the country. Rural areas (in particular Eden and Penrith) are highlighted as having cases driven by external force of infection, though themselves are less likely to sustain ongoing transmission at the local level (being blue in the Within-region map). Time-series plots of the three model components are shown in Figure S1 for 4 arbitrarily chosen LTLAs.

**Figure 4:**
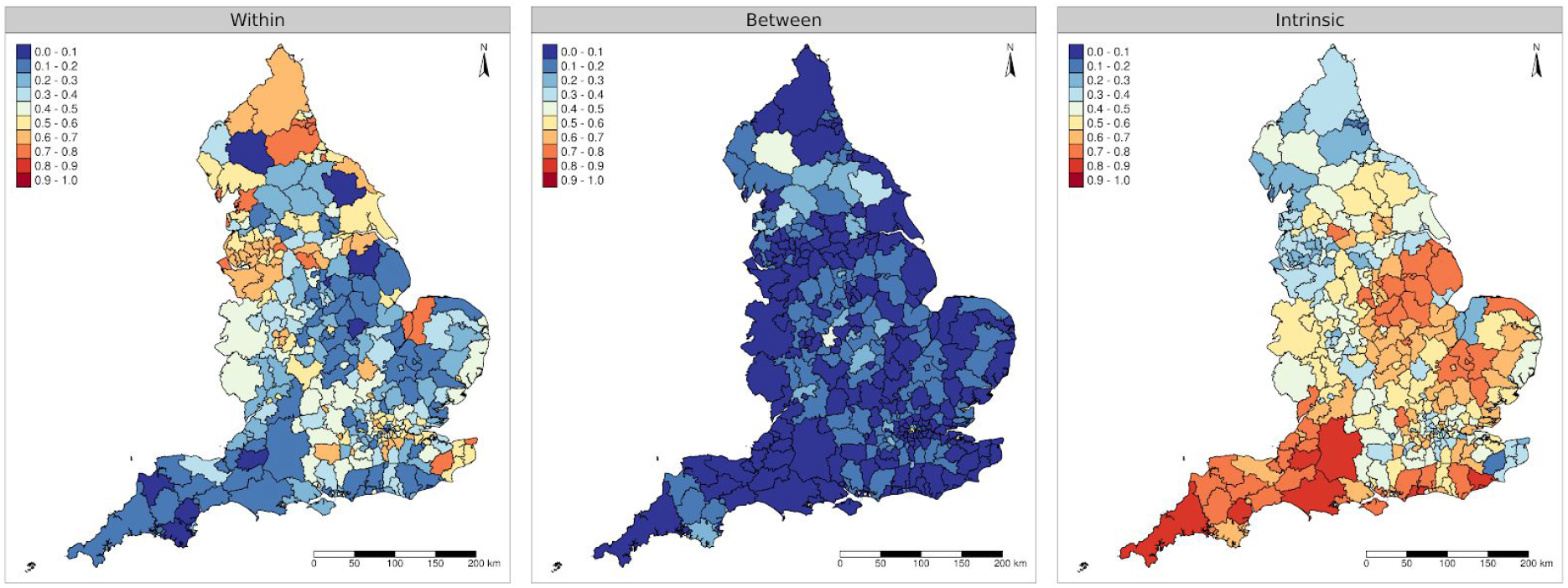
Contribution of Within- and Between-LTLA and intrinsic infection rate as a proportion of overall incidence.

Figure 5 (left) shows a choropleth map of the region-level autoregressive component random effect. As with the Within-region component map in Figure 4, this may be interpreted as a region-specific propensity to generate more cases given the current number of infected individuals, and is analogous (but not equal!) to a region-specific reproduction number. There is evidence for a spatially diverse set of regions having a higher daily increase of cases than others, having accounted for current case numbers, population size, and human mobility. South and West Cumbria leads these regions, with unusually high within-region incidence. Figure 5 (right) shows the standard error associated with each estimate of the random effect, providing a quantification of uncertainty about the disease process in each LTLA. There are clear regions of the country where uncertainty is markedly higher than elsewhere, indicating an equality in our current understanding of the disease process.

**Figure 5:**
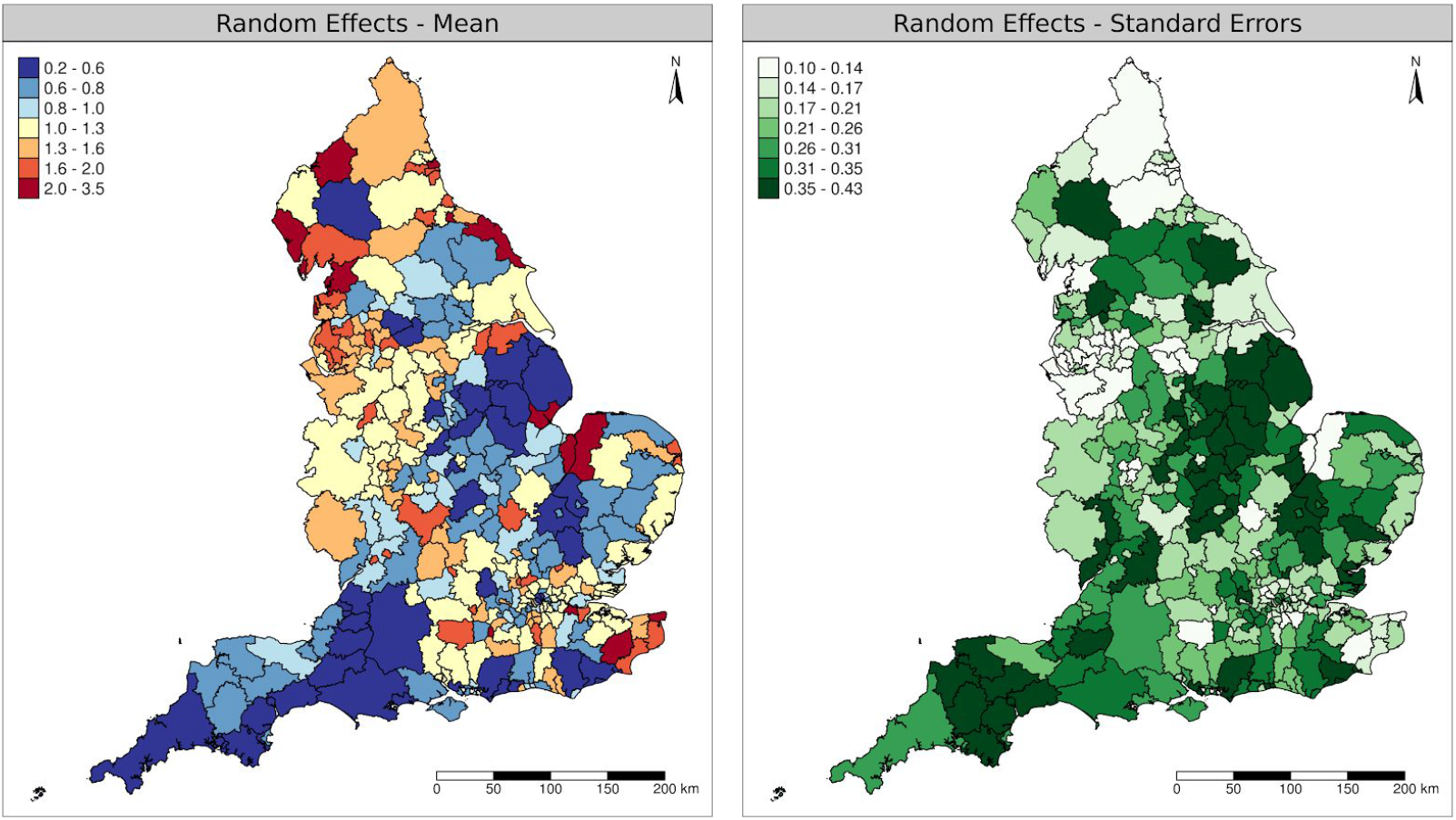
Left: Estimated district specific deviations α_*i*_ − λ_0_ expressed as a Relative Risk (1 represents national average). Higher values represent regions with a greater propensity for self-sustained disease transmission. Right: standard error of Within-LTLA random effect providing a measure of uncertainty about the disease process within each region.

Predictive time series plots are shown in Supplementary Figure S2, indicating a good quality of model fit.

## 4. Discussion

In this paper, we have applied a spatiotemporal stochastic process model to investigate the dynamics of COVID-19 positive tests reported by PHE at the LTLA level across England. Though phenomenological in nature, the model demonstrates its ability to capture a high level of stochastic variation within and between discrete regions, with our results showing that the incidence of COVID-19 infections, and the dynamics of the disease process causing infections, varies widely across the UK. By decomposing incidence particularly into Within- and Between-region components (Figure 4), **we show that the COVID-19 epidemic in England is still highly volatile, with regional activity that is not apparent when looking at aggregated national data (cf. Figure 1)**.

Our results indicate a national decline in the incidence of reported positive COVID-19 cases, which in the absence of changes to BSI and population behaviour more widely may be expected to continue. However, should adherence to the BSI in place decline further, or governmental relaxations be introduced, we would expect this to reverse. Careful and detailed spatial monitoring of the impact of reversal on cases should be undertaken, which if coupled to analytic procedures such as ours can distinguish between regions affected in a heterogeneous way. **The analysis demonstrates considerable localisation of ongoing transmission across England; the spatial granularity with which public health interventions are applied should reflect this**.

COVID-19 testing resources, even for resource-rich countries such as the UK, will always be constrained by cost and availability. Nevertheless, analytic methods such as ours may be used to direct testing resources to regions of greatest uncertainty. This method has been described by Kabaghe *et al*. (2017), for use in resource-poor settings, though is equally applicable here. The random effect standard errors in Figure 5 (right) may be used to **coordinate and target surveillance in an adaptive fashion across the country, focusing on areas with higher uncertainty** (high standard error). Surveillance information such as this may then be fed back into analysis to improve future estimates: testing resources are therefore directed to areas of poor knowledge, rather than being used needlessly in regions where the pattern of disease is already well understood.

It is clear that more needs to be done to improve the spatial granularity of official case data reporting to incorporate fine-scaled demographic and health variation, and the need to report negative tests alongside those positive in order to be able to make statements on certainty of prevalence estimates. As described below, projects such as the “COVID Symptom Study” clearly show the advantage of finely resolved spatial “prevalence” data, though lack the rigour of case definition afforded by clinical testing.

## Limitations

Our model has a number of limitations due to the scope of the underlying case data, measures of commuting, and geographical extent. Currently, we only consider England due to the availability of spatially resolved data at a fine scale, though as the other three nations of the UK improve their data resolution our method will naturally extend to them. We do not currently account for cross-border migration, hence we expect to see significant “edge effects”, particularly in LTLAs on the border with Scotland and Wales.

As mobility/interaction patterns were based on work-based commuting census information scaled by private vehicle usage, the model may overestimate the between-region effect for locations with large proportions of non-working individuals, and may not represent highly urban areas where car use is lower than the national average. We investigated this by adding a random effect to the Between-region model component, though this did not improve the fit of our model to the data significantly.

The biggest limitation of our analysis is the reliability of the under-pinning COVID-19 test positive data from PHE. These data were used to demonstrate a pragmatic, fast, and informative analytic technique, addressing our aims of highlighting “hotspots” of epidemic activity and providing a method of addressing spatially-varying measures of uncertainty for targeted surveillance. PHE has recently released LTLA-level data publicly, providing an opportunity to more fully exploit spatial analytic powers. Additionally, several factors that have been shown to contribute to risk from COVID-19 have been identified, including age, comorbidities, and possibly social deprivation and ethnicity. These factors show marked variation at small spatial scales, for example within a single town, and higher resolution case data may allow us to account for these variables to improve our predictions. As an example, **Figure S3 shows the GB-wide geographical variation, at Lower Super Output Area (LSOA)-level spatial resolution, in the rate of positive symptom reporting by users of the COVID-19 Symptom Tracker App developed by King’s College London and ZOE (https://covid.joinzoe.com/) in the 14 days to 16 April 2020**. Local symptom incidence shows a five-fold variation (upper left panel), albeit with relatively wide lower 5% and upper 95% probability limits (lower panels). Though data from the App provide only a crude estimate of number of COVID-19 cases, their use demonstrates a critical feature missing from current data streams: **information is available not only on symptom positive individuals, but also on symptom negative individuals**, allowing estimates of spatial prevalence with well characterised uncertainty (Diggle and Giorgi, 2019, p85).

## Data Availability

Data used in this paper are freely available from UK government sources, with the exception of the traffic volume data which is available from the UK Department for Transport on request.

https://coronavirus.data.gov.uk

https://ons.gov.uk

https://www.gov.uk/government/organisations/department-for-transport

## Supplementary Figures and Tables

**Figure S1:**
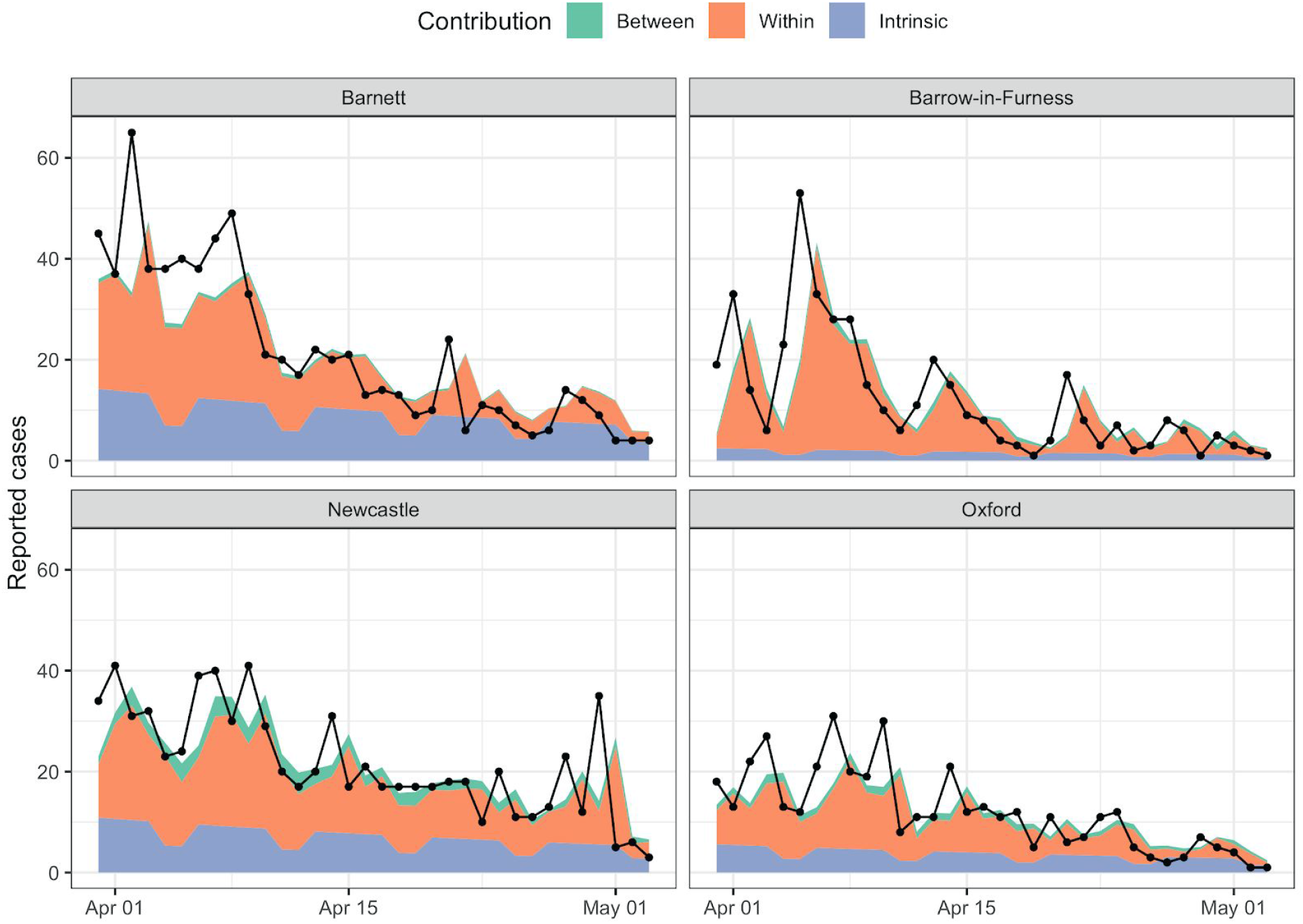
Contributions to overall transmission from Intrinsic, Within-, and Between-region transmission for 4 LTLAs in England.

**Figure S2:**
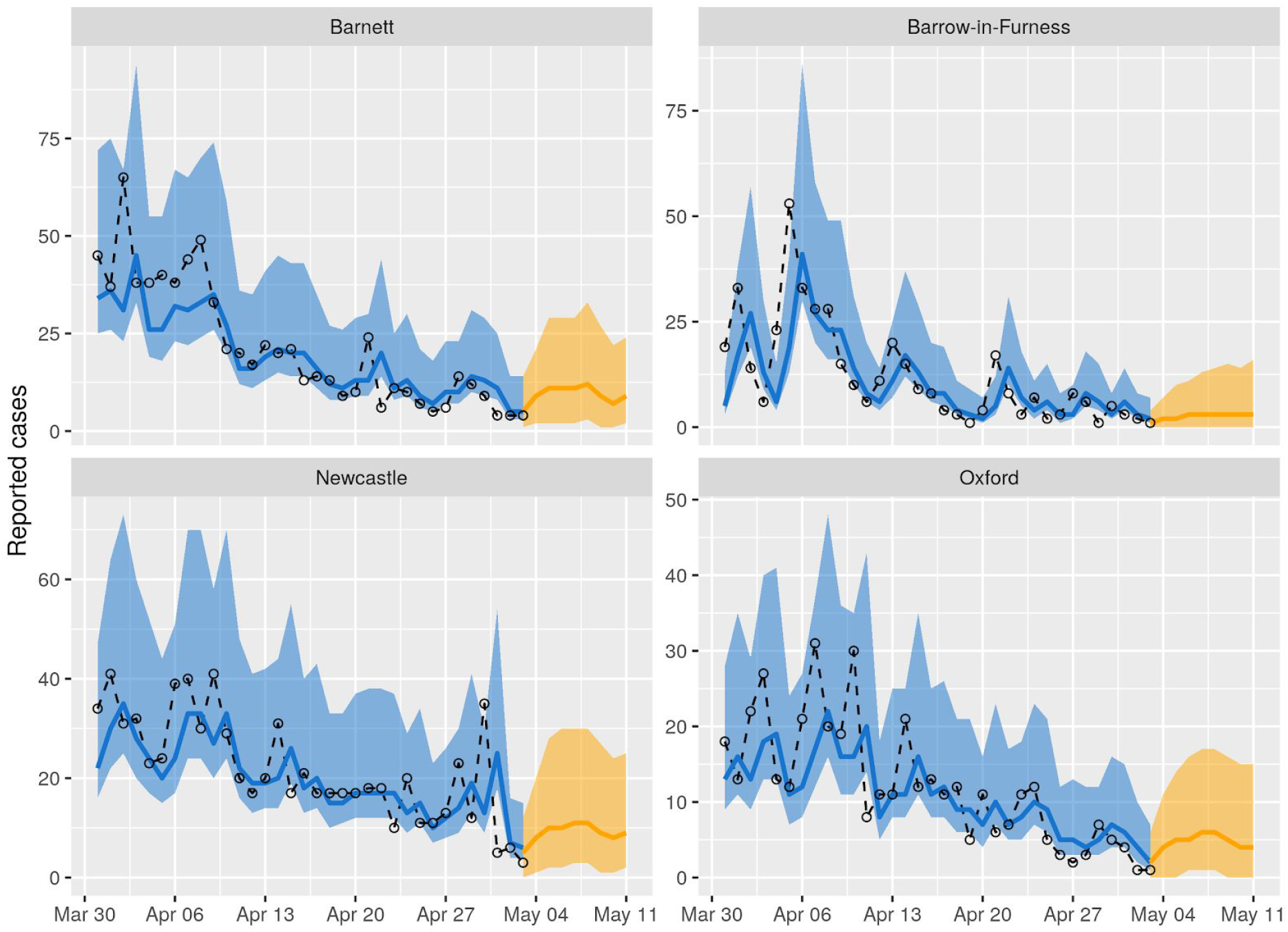
Predictive plots of case time series for 4 arbitrarily chosen LTLAs, demonstrating the fit of the spatiotemporal stochastic time series model. Observed cases are indicated by the black dashed line and dots, the median in-sample prediction by the solid blue line (95% confidence interval by the blue shaded region), and 7 day forward prediction similarly in orange.

**Figure S3:**
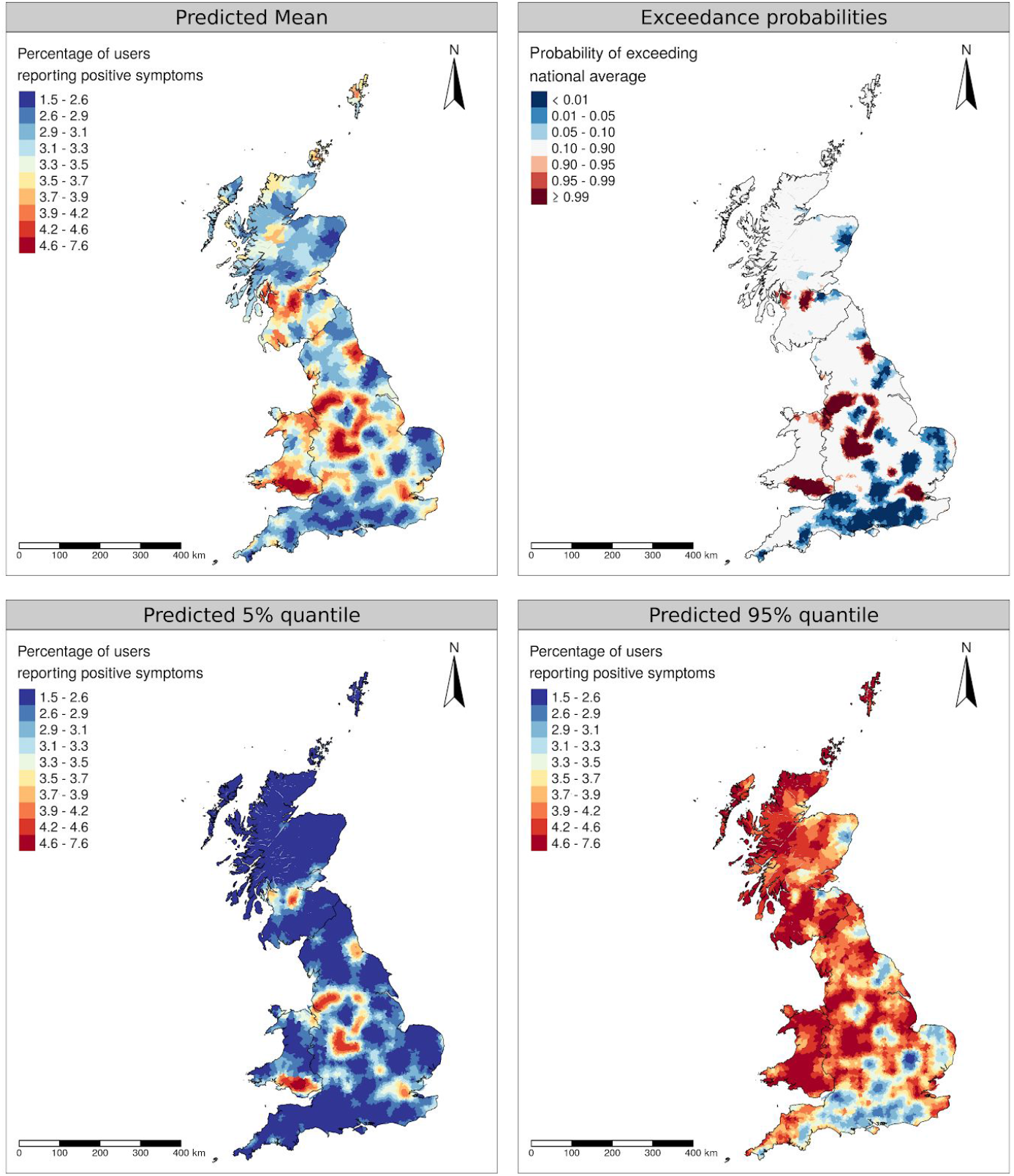
Geographical variation in the proportion of active COVID-19 Symptom Tracker App users reporting positive symptoms at least once over the 14-day period to 16 April 2020. Point estimate of prevalence (upper left panel), probability that underlying prevalence is greater than the GB-wide average (upper right panel), 5% lower and 95% upper probability limits (lower panels).

**Table S1:** Parameter estimates for the spatiotemporal time-series model for COVID-19 in England LTLAs. ^1^Absolute incidence; ^2^Relative Risk; ^3^Natural scale.

| Model component | Variable | Relative Risk Estimate (95% CI) |
| --- | --- | --- |
| Baseline | Intercept $\varepsilon_0$ | 7926 (6651, 9447)<sup>1</sup> |
| | Linear time, $\delta_1$ | 0.978 (0.976, 0.980)<sup>2</sup> |
| | Weekend, $\delta_2$ | 0.536 (0.502, 0.572)<sup>2</sup> |
| Within-region | Intercept $\lambda_0$ | 4.11 (1.96, 8.62)<sup>1</sup> |
| | log Population $\beta_1$ | 1.53 (1.35, 1.74)<sup>2</sup> |
| | Random effect variance $\tau^2$ | 0.25<sup>3</sup> |
| Between-region | Intercept $\phi_0$ | 3.00e-4 (6.35e-5, 1.42e-3)<sup>1</sup> |
| | log Population $\gamma_1$ | 0.460 (0.368, 0.575)<sup>2</sup> |
| | log Commuting $\gamma_2$ | 0.387 (0.198, 0.757)<sup>2</sup> |
| Observation model | over-dispersion $r$ | 0.158 (0.149, 0.166)<sup>3</sup> |

